# Block-Level Deep Sleep Consolidation and Environmental Modifiers in Long COVID with Dysautonomia: A Longitudinal N-of-1 Wearable Study

**DOI:** 10.64898/2026.08.06.26359860

**Authors:** Michael F. Hammer

**Affiliations:** BIO5 Institute, University of Arizona, Tucson AZ 85821

**Keywords:** Long-covid, autonomic nervous system, sleep disorder, sleep hygiene, sleep architecture, viral illness, space weather

## Abstract

**Background:** Sleep architecture fragmentation is common in Long COVID and dysautonomia, yet the relationship between block-level deep sleep consolidation and next-day functional wellbeing has not been characterized longitudinally in this population. We tested the hypothesis that block-level deep sleep architecture predicts next-day wellness better than aggregate stage duration.

**Methods:** A prospective N-of-1 longitudinal study was conducted across 78 nights between January 1 and March 25, 2026 using a consumer wearable (Garmin Index Sleep Monitor). Next-day wellbeing was assessed using the Overall Feeling Score (OFS; 0–10 scale). A five-tier Architecture Group (AG) classification operationalized consolidation quality. A custom predictive model (Pendulum v5.2) quantified consolidation through block-level weighting and contextual penalties. Space weather metrics (Kp index, daily electron fluence, Load Score) were correlated with sleep architecture and OFS across same-day and lagged alignments. A secondary hierarchical regression examined illness as an independent covariate across the full dataset.

**Results:** Architecture Group explained 56.7% of next-day OFS variance (r = 0.753, p < 0.001), compared with 17.0% for total deep sleep duration (r = 0.412) and 4.8% for the Garmin Sleep Score (r = 0.220). Pendulum v5.2 explained 43.0% of variance (r = 0.656). The deep sleep fragmentation phenotype occurred on 25.6% of nights despite adequate total deep sleep. Space weather explained 14.2% of OFS variance through architecture degradation; Load Score at 1-day lag was the strongest space weather predictor (r = −0.328, p = 0.004). A binary illness indicator explained an additional 5.7% of OFS variance beyond architecture and space weather (full-model R² = 55.2%), with a mean illness-night residual of −0.56 (SD = 0.36; 95% CI [−0.72, −0.40]), consistent with immune/viral flares reducing OFS through pathways independent of sleep architecture.

**Conclusions:** Block-level deep sleep consolidation quality is a substantially stronger predictor of next-day wellbeing than total stage duration in Long COVID with dysautonomia. Space weather constitutes a measurable environmental modifier operating through architecture degradation. Immune and viral flares constitute a significant architecture-independent confound consistent with direct neuroinflammatory effects on functional capacity. This framework may be applicable across conditions where sleep architecture fragmentation plays a pathophysiological role.

**Author Summary:** Sleep fragmentation is common in Long COVID and dysautonomia, but the relationship between block-level deep sleep consolidation and next-day functional outcome has not been characterized longitudinally. Consumer wearables can capture block-level architecture at ecological scale, yet commercial sleep scores show poor predictive validity in this population. This study demonstrates that deep sleep *consolidation quality*—not total stage duration—is the dominant predictor of next-day wellbeing in Long COVID with dysautonomia (Architecture Group r = 0.753, explaining 56.7% of variance). Space weather constitutes a measurable environmental modifier operating through architecture degradation via dissociable Kp and electron fluence pathways. The block-level framework and Pendulum Model may generalize to other conditions where ANS dysregulation impairs sleep stage consolidation.

## 1. INTRODUCTION

The post-acute sequelae of SARS-CoV-2 infection (PASC), commonly referred to as Long COVID, represents one of the most clinically consequential and mechanistically complex challenges to emerge from the COVID-19 pandemic. Prevalence estimates vary with case definition and follow-up duration, but large-scale studies suggest that 10–30% of individuals who contract SARS-CoV-2 develop persistent symptoms lasting beyond 12 weeks post-infection [1]. Symptoms span multiple organ systems and include post-exertional malaise, cognitive dysfunction, dysautonomia, respiratory impairment, and profound sleep disturbance, among others [2, 3]. The heterogeneity of Long COVID presentations has complicated both clinical management and research design, with different patient subgroups exhibiting distinct pathophysiological profiles that do not respond uniformly to standardized interventions.

Sleep disturbance is among the most consistently reported Long COVID symptom, with estimates ranging from 25 to 65% of patients describing non-restorative sleep, insomnia, or hypersomnia in post-acute surveys [4]. Despite its high prevalence, the mechanisms underlying Long COVID-associated sleep disruption remain poorly characterized [5]. Most existing literature describes sleep disturbance in terms of subjective complaint scales or total sleep time estimates from consumer wearables, without characterizing the architectural quality of sleep, the temporal distribution, block-level consolidation, and stage sequencing that determine whether time spent asleep translates into physiological restoration. As a result, the specific pathways through which Long COVID disrupts sleep and impairs next-day function remain largely unmapped at the architecture level.

Consumer wearable devices capable of continuous sleep monitoring represent a transformative opportunity for longitudinal sleep research. In contrast to polysomnography (PSG), which captures single or limited-duration laboratory nights under artificial conditions, consumer wearables enable naturalistic, ecologically valid sleep data collection across weeks to months at no per-night cost. Validation studies report moderate PSG agreement for broad stage categories in healthy adults [6, 7].

Single-participant longitudinal research designs have a long and productive history in clinical science, enabling the characterization of within-person physiological dynamics at a resolution that population studies cannot achieve [8]. In sleep medicine, n = 1 studies have contributed foundational observations in chronobiology, insomnia treatment, and circadian rhythm disorders, where individual variability is large relative to population means and within-person replication across nights provides robust internal validity [9]. The present study employs this design to characterize, with high temporal resolution, the extent to which sleep quality contributes to

Long-COVID-related dysfunction, the mechanisms through which Long COVID-related dysautonomia disrupt sleep consolidation, and to develop and validate a predictive model relating block-level sleep architecture to next-day functional outcome. The primary objective was to determine whether block-level consolidation quality predicts next-day wellness more accurately than aggregate sleep stage duration, and to identify the ANS-level and environmental determinants of architectural variation across a three-month observational period.

## 3. METHODS

### 3.1 Study Design

This N= 1 prospective, single-participant, longitudinal observational case study was conducted from January 1 to March 25, 2026, encompassing 84 nights of habitual sleep recorded at the participant’s primary residence. A prior baseline period (November 1, 2025 onward) informed protocol development but was excluded from primary correlational analyses. The finalized “clean protocol”—a standardized set of behavioral, dietary, pharmacological, and environmental conditions—was developed empirically during baseline and maintained across study nights (details in **Supplementary Section I** and **Table S1a**). The participant’s stable nightly medication and supplement protocol, established through empirical optimization during the baseline period, is described in detail in **(Table S1b**). Sympathomimetic agents (pseudoephedrine, albuterol) were used on isolated occasions and constitute confound events requiring asterisking of affected nights.

Data collection, model development, and analysis proceeded iteratively, with prospectively documented refinements applied retroactively to the full dataset for validation, consistent with established n=1 self-experimentation approaches. Primary analyses were restricted to “clean” nights (n = 78), defined as nights without prespecified confounds. See **Supplementary Section II** for Nights description of exclusion criteria and reasons for asterisked nights. Twelve nights were flagged, with six excluded from the primary analysis window (**Table S2**). Data collection continued beyond the primary analysis period through April 24, 2026 (30 additional nights). These extended-period data are not included in primary correlational analyses but are documented in **Supplementary Table S3** and referenced where they provide observational context for Discussion findings.

### 3.2 Participant

The participant is a 71-year-old male with a diagnosis of Long COVID (post-acute sequelae of SARS-CoV-2 infection, PASC), initially acquired in January 2022. Within six months after initial recovery from the primary infection and 5-day hospitalization, a series of novel symptoms evolved. Major sequelae relevant to sleep and the present study include: (1) autonomic nervous system dysfunction (dysautonomia), presenting with orthostatic intolerance, heart rate variability dysregulation, and gastrointestinal dysmotility; (2) shortness of breath, characterized by diaphragmatic weakness with a shift to voluntary respiratory control under cognitive or physical load; (3) sleep-disordered breathing; and (4) neurological sequelae including occasional malaise, vestibular dysfunction, and migraine susceptibility. The participant maintained full-time professional employment throughout the study period. Overnight supplemental oxygen was initiated in October 2025 following documentation of nocturnal desaturations (SpO₂ nadirs in the mid-80s%) on room air, confirmed by wearable pulse oximetry. At the time of primary study enrollment, the participant had been established on 2 L/min supplemental O₂ via nasal cannula as the sole respiratory intervention (**Table S1a**). Documentation of sleep study results are presented in **Supplementary Section III**.

### 3.3 Wearable Device and Data Collection

The participant wore a Garmin Index Sleep Monitor L/XL (model 010-03024-00) continuously during sleep throughout the study period. Sensor placement was standardized to the inside of the left bicep, which the participant had empirically established as providing more reliable optical contact during side-sleeping (the habitual sleep position) than the standard dorsal-wrist placement. Nightly metrics extracted from Garmin Connect included sleep stage durations, block-level deep and REM architecture (start times, durations, and sequence), HRV, SpO₂ (average and nadir), respiration rate, restless moments, proprietary Stress and Sleep scores, and breathing variation events; the full metric list is provided in **Table S3**. Nightly sleep architecture (e.g., awake events, N3 deep and REM stages) was manually extracted from the Garmin Connect visualization.

### 3.4 Outcome Measure: Overall Feeling Score (OFS)

The primary outcome variable was a self-reported Overall Feeling Score (OFS), recorded each day after a minimum two-hour post-waking period. OFS was recorded on a continuous 0–10 scale anchored at 0 (complete functional incapacity, maximal symptom burden) and 10 (self-reported functional baseline prior to Long COVID onset), with the active range of variability observed across the study period falling between approximately 2 and 9. The OFS captures integrated next-day wellness, incorporating cognitive clarity, physical energy, vestibular stability, and absence of symptom burden. OFS was reported after a minimum of a two-hour post-waking window to mitigate confounding from immediate post-waking grogginess. The OFS was reported without knowledge of the preceding night’s biometric data.

### 3.5 Sleep Quality Scoring: The Pendulum Model (v5.1/v5.2)

A custom predictive model, designated Pendulum v5.1 and refined to v5.2, was developed iteratively over the study period to translate block-level sleep architecture into a predicted feeling score (OPF). The model was designed to test the hypothesis that consolidation quality, operationalized as the quadratic distribution of deep and REM sleep time across blocks weighted by peak block duration, predicts next-day wellness better than aggregate stage totals alone. Full algorithm specifications, including the Deep Score, REM Score, Sleep Quality Score (SQS), best-block multiplier schedule, N-attempts penalty, four-night bank bonus, Deep:REM ratio penalty, and context adjustments, are provided in **Supplementary Section IV**.

### 3.6 Sleep Architecture Pattern Classification

A categorical architecture pattern classification system (Architecture Group, rated on a 1–5 scale) was developed and applied to each night to complement the continuous OPF metric. The system operationalizes the fragmentation phenotype at the level of clinical interpretation, distinguishing architectures by first-block duration and total block count. The five-tier classification is described in **Table 1**. Details on block length measurement and categorization of REM block patterns are given in **Supplementary Section V**.

**Table 1.** Architecture Group Classification.

| Tier | Label | Criteria | Interpretation |
| --- | --- | --- | --- |
| 1 | AG1 | First block <40 min AND/OR total deep <50 min | Insufficient homeostatic discharge; likely absent glymphatic clearance |
| 2 | AG2 | ≥5 blocks; first block <40 min AND total ≤100 min | Classic stutter phenotype; N-attempts penalty fires |
| 3 | AG3 | 1–4 blocks; first block 40–49 min | Near-threshold consolidation; borderline glymphatic clearance |
| 4 | AG4 | ≥5 blocks; first block ≥40 min OR total deep >100 min | Fragmented but meaningful consolidation achieved; high-volume stutter |
| 5 | AG5 | 1–4 blocks; first block ≥50 min | Optimal consolidation; full or near-full glymphatic clearance |
| *Combined-block convention: Two consecutive deep blocks separated by <20 min light sleep (with no awake events) are combined into a single block for group assignment purposes only; this convention does not affect SQS or OPF calculations.* |  |  |  |
| *REM subclassification appended where clinically relevant: 'a' (≥75 min, adequate); 'b' (60–74 min, suboptimal); 'c' (<60 min, crash state). Primary analyses use full dataset; sensitivity analyses restricted to REM-a nights showed AG means within 0.02–0.39 OFS points of full-sample values.* |  |  |  |

### 3.7 Environmental Variables: Space Weather Metrics

Three space weather metrics were recorded nightly and linked to sleep and OFS data for temporal correlation analysis: Kp Index (daily peak), 2MeV Electron Flux Load Score (daily), and Daily Electron Fluence. For a description of these indices, consult **Supplementary Section VI.** All values were sourced from NOAA’s Space Weather Prediction Center (NOAA SWPC; https://www.swpc.noaa.gov). Prior-day confirmed values were used (i.e., values logged the morning following the sleep night) as NOAA finalizes electron fluence data at end of UTC day.

### 3.8 Statistical Analysis

All correlations reported are bivariate Pearson product-moment correlation coefficients (r). Statistical significance was assessed using two-tailed t-tests against the null hypothesis r = 0, with significance threshold α = 0.05. No corrections for multiple comparisons were applied to the primary model performance correlation; exploratory correlation analyses (space weather metrics, raw architecture variables) are reported with p-values and interpreted in the context of the number of tests conducted. Effect sizes are reported as r² (proportion of variance explained). Mean absolute error (MAE) was calculated as the average absolute difference between OPF and OFS across nights. Space weather correlations were analyzed across three temporal alignments: same-day, one-day lag, and two-day lag. All analyses were conducted in Microsoft Excel (version 16.107.1) using built-in Pearson correlation and t-distribution functions. The iterative model development approach introduces a degree of circularity in model performance estimates, as model parameters were refined using the same dataset against which final performance is reported; this limitation is standard in n = 1 model development work and is addressed in the Discussion.

## 4. RESULTS

### 4.1 Dataset Overview

Some of the key results of the study are presented in **Table 2**. The mean deep sleep was 71.8 ± 25.0 minutes (range 35–146 min), Mean REM was 97.1 ± 19.0 minutes (range 30–128 min), mean OFS was 4.9 ± 1.8 (range 0.5–8.0), and mean overnight HRV was 34.3 ± 9.9 ms. Despite support with 2L O2 via cannula, the mean SpO₂ nadir was 92.8 ± 2.7%, reflecting occasional failure in delivery of supplemental oxygen. REM category ‘a” dominated (n = 62, 79.5%) with categories ‘b’ and ‘c’ only accounting for 14.1%), and 6.4% of nights. Sensitivity analyses restricted to REM-a nights produced Architecture Group OFS means within 0.39 points of full-sample values (largest deviation AG2, n = 3 b/c nights); all primary analyses use the full 78-night clean dataset.

**Table 2.** Summary of Study Dataset.

| Variable | Mean ± SD | Range |
| --- | --- | --- |
| **Sleep Architecture** |  |  |
| Deep sleep (min) | $71.8 \pm 25.0$ | 35–146 |
| REM sleep (min) | $97.1 \pm 19.0$ | 30–128 |
| Light sleep (min) |  |  |
| Total sleep (min) |  |  |
| Awake events (n) |  |  |
| Deep:REM ratio |  |  |
| Architecture Group (1–5) |  | 1–5 |
| **Physiological Monitoring** |  |  |
| HRV overnight (ms) | 34.3 ± 9.9 |  |
| SpO<sub>2</sub> nadir (%) | 92.8 ± 2.7 |  |
| Garmin Sleep Score |  |  |
| **Outcome Variable** |  |  |
| OFS (Overall Feeling Score) | 4.9 ± 1.8 | 0.5–8.0 |
| **REM Category Distribution** |  |  |
| REM group 'a' (≥75 min, adequate) | n = 62 (79.5%) |  |
| REM group 'b' (60–74 min, suboptimal) | n = 11 (14.1%) |  |
| REM group 'c' (<60 min, crash state) | n = 5 (6.4%) |  |
| **Space Weather (primary study period)** |  |  |
| Kp Peak (geomagnetic index) |  | 0–9 |
| Daily electron fluence (×10<sup>8</sup> e/cm<sup>2</sup>/sr/day) |  |  |
| Load Score (cumulative flux) |  | 0–5+ |

Cells with missing Mean ± SD values were not computed as primary summary statistics; raw values available in full dataset (Supplementary Data). OFS = Overall Feeling Score (rescaled 0–10 instrument; see Methods 3.4).

### 4.2 Sleep Architecture Characteristics

**Table 3** lists the distribution of Architecture Groups: AG1 (n = 17, 21.8%), AG2 (n = 12, 15.4%), AG3 (n = 27, 34.6%), AG4 (n = 8, 10.3%), and AG5 (n = 14, 17.9%). AG3 was the modal architecture, reflecting a pattern of moderate consolidation achieved on more than one-third of nights. The combined prevalence of fragmented “stutter” architectures (AG2 and AG4) was 25.6% of clean nights, occurring with less than half the frequency of well-consolidated architectures (AG3 and AG5, n = 41 nights, 52.6%). **Figure 1** illustrates the distribution of OFS outcomes associated with each AG for nights with REM pattern ‘a’. The relationship between AG and next-day OFS was monotonically increasing across all five groups (**Table 3**). Mean OFS for REM pattern ‘a’ nights ranged from 2.85 ± 1.15 in AG1 to 7.04 ± 0.58 in AG5, representing a 4.19-point absolute range across the architectural quality spectrum.

**Figure 1.**
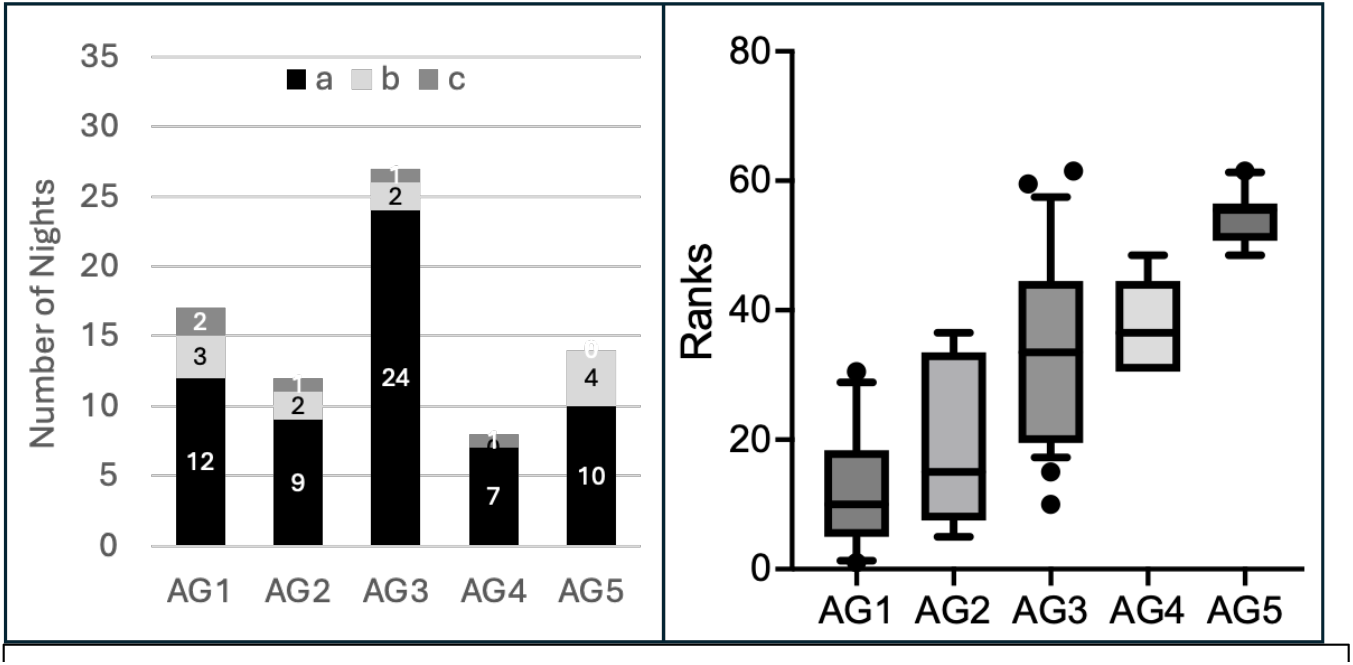
Architecture Group Distribution. **Left Panel:** Bar chart frequency by architecture group (AG) and numbers of nights with REM patterns “a”, “b” and “c” (n=78). **Right Panel:** Kruskal-Wallis rank distribution of OFS by AG. The y-axis represents ranked OFS values as computed for non-parametric group comparison. The monotonic increase in rank distribution across AG1-AG5 confirms the ordinal relationship between consolidation quality and next-day wellness (K-W statistic= 36.57, p<0.0001).

**Table 3.** Architecture Group Characteristics and OFS Outcomes (n=78 clean nights)

| Architectural Group (AG) | n (%) | Mean Deep (min ± SD) | Mean OFS (all nights ± SD) | Mean OFS (REM-a only ± SD) | REM-a n |
| --- | --- | --- | --- | --- | --- |
| 1 | 17 (21.8%) | 51.5 ± 16.1 | 2.76 ± 1.15 | 2.85 ± 1.32 | 12 |
| 2 | 12 (15.4%) | 81.6 ± 17.7 | 4.23 ± 1.49 | 3.83 ± 1.43 | 9 |
| 3 | 27 (34.6%) | 60.0 ± 14.6 | 5.19 ± 1.39 | 5.33 ± 1.34 | 24 |
| 4 | 8 (10.3%) | 106.8 ± 19.7 | 5.75 ± 0.56 | 5.73 ± 0.60 | 7 |
| 5 | 14 (17.9%) | 90.7 ± 20.7 | 6.80 ± 0.58 | 7.04 ± 0.44 | 10 |

All values computed from CSV (n = 78 clean nights, Jan 1–Mar 25, 2026, excluding 1/28/26). REM-a = nights with ≥75 min total REM. AG4 has the highest mean total deep sleep of any group (106.8 ± 19.7 min), illustrating the dissociation between deep sleep quantity and consolidation quality central to this study's thesis (see Discussion 5.2).

Deep sleep total minutes showed substantial night-to-night variability (range 35–146 min, CV 34.8%) not fully captured by AG alone (**Table S3**), reflecting the distinction between total duration and consolidation quality. Notably, the longest deep sleep night in the dataset (March 11, 2026: 146 minutes, asterisked) was associated with an OFS of only 1.4 due to pharmacological REM suppression by pseudoephedrine, producing a severely inverted

### 4.3 Model Performance: Pendulum v5.1 and v5.2

The Pendulum v5.1 model achieved a Pearson r of **0.638** (p < 0.001) between predicted feeling score (OPF v5.1) and actual OFS across the 78-night clean dataset, explaining **40.7%** of OFS variance (**Table S5**). The OPF v5.2 model achieved r = **0.656** (p < 0.001), explaining **43.0%** of variance. Both represent substantial improvements over the original linear V3 model (r = 0.533, 28.4% variance explained, clean n = 46), with each structural addition contributing measurable predictive gain (Table 4). Mean absolute error on the clean dataset was **0.374** OFS units for v5.1 and **0.372** for v5.2 (raw OFS scale). **Figure 2** shows the distribution of predicted versus actual OFS values across clean nights, color-coded by Architecture Group. The model correctly identified the full range of OFS outcomes without systematic bias: mean residual was approximately +0.02 OFS units across the clean dataset.

**Figure 2.**
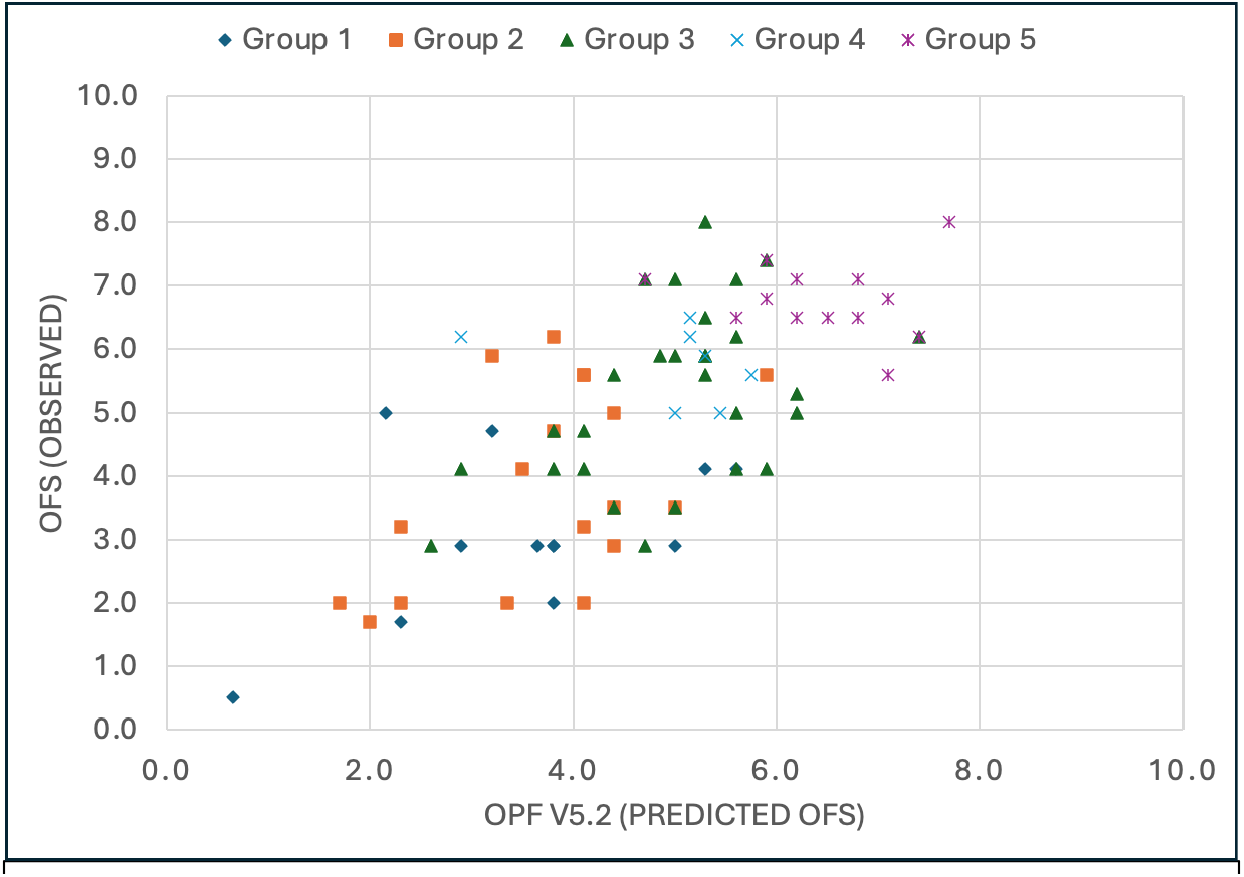
OPF v5.2 (predicted vs OFS scatter plot: n=78 clean nights, color-coded by Architecture Group 1–5. r=0.656, p<0.001.

**Table 4.** Competing Predictors of OFS: Head-to-Head Comparison (clean n=78)

| Predictor | r | p | r<sup>2</sup> |
| --- | --- | --- | --- |
| **Sleep Architecture Predictors** |  |  |  |
| Architecture Group (categorical) | 0.753 | <0.001 | 56.7% |
| OPF v5.2 (predicted OFS) | 0.656 | <0.001 | 43.0% |
| OPF v5.1 | 0.638 | <0.001 | 40.7% |
| Longest Deep Block (min) | 0.488 | <0.001 | 23.8% |
| Deep Score | 0.484 | <0.001 | 23.4% |
| SQS v5 (Sleep Quality Score) | 0.464 | <0.001 | 21.5% |
| Deep min total | 0.412 | <0.001 | 17.0% |
| Deep Wt Avg Block (min) | 0.370 | <0.001 | 13.7% |
| **Comparator Predictor** |  |  |  |
| Garmin Sleep Score (proprietary) | 0.220 | 0.053 | 4.8% |
| REM min total | 0.166 | 0.146 | 2.8% |
| **Space Weather** |  |  |  |
| Kp Peak (geomagnetic index) | -0.249 | 0.028 | 6.2% |

† Longest Deep Block n = 66; 12 nights with non-parseable single-block entries excluded from this correlation. All other predictors n = 78. Ordered by descending r within section. ns = not significant (p > 0.05).

**Table 4** presents a head-to-head comparison of competing predictors. Architecture Group was the single strongest predictor (r = **0.753**), outperforming OPF v5.2 (r = **0.656**) and OPF v5.1 (r = **0.638**). The most clinically relevant comparison is against the Garmin proprietary Sleep Score (r = **0.220**, p = **0.053**), which explained only **4.8%** of OFS variance compared to **56.7%** for Architecture Group and **43.0%** for OPF v5.2. Total deep sleep minutes (r = **0.412**) outperformed the Garmin score but underperformed all consolidation-weighted metrics, confirming that block-level quality adds substantial predictive information beyond total stage duration.

### 4.4 The Deep Sleep Fragmentation Phenotype

The fragmentation phenotype, characterized by multiple short deep sleep blocks with a best block below 40 minutes (AG2 and AG4), occurred on **25.6%** of clean nights (n = 20). Critically, this phenotype was mechanistically distinct from simple deep sleep reduction: many fragmented nights achieved adequate or high total deep sleep (mean deep minutes for AG2 nights: **81.6 ± 17.7** min; AG4: **106.8 ± 19.7** min), yet produced systematically worse OFS outcomes than consolidated nights with comparable or lower total deep sleep.

**Figure 3** illustrates this contrast using two exemplar nights selected to minimize environmental confounds while maximizing architectural contrast. February 25, 2026 (AG2) produced 78 minutes of deep sleep across **five** blocks (combined-block convention applied; longest effective block **39** minutes), with an OFS of 2.9. March 9, 2026 (AG5) produced 80 total minutes of deep sleep in two blocks—a single 69-minute first block followed by a 12-minute secondary block— with an OFS of 6.8. The 3.9-point OFS differential emerged from near-identical total deep sleep, with the consolidated night achieving glymphatic-clearance-sufficient block durations (69 minutes; see Discussion, **Section 5.2**) while the fragmented night failed to sustain any block beyond 39 minutes. Environmental conditions: February 25 (Kp 3.8, Load Score 4, daily fluence 4.4 × 10⁻⁸; mild environmental headwind); March 9 (Kp 2.2, Load Score 2, daily fluence 1.0 × 10⁻⁸; clean). The February 25 environmental context is noted as a caveat; the Load Score 4 on that night did not trigger the context adjustment threshold for model penalties, and the architectural fragmentation was consistent with other fragmented nights under similar or better environmental conditions.

**Figure 3.**
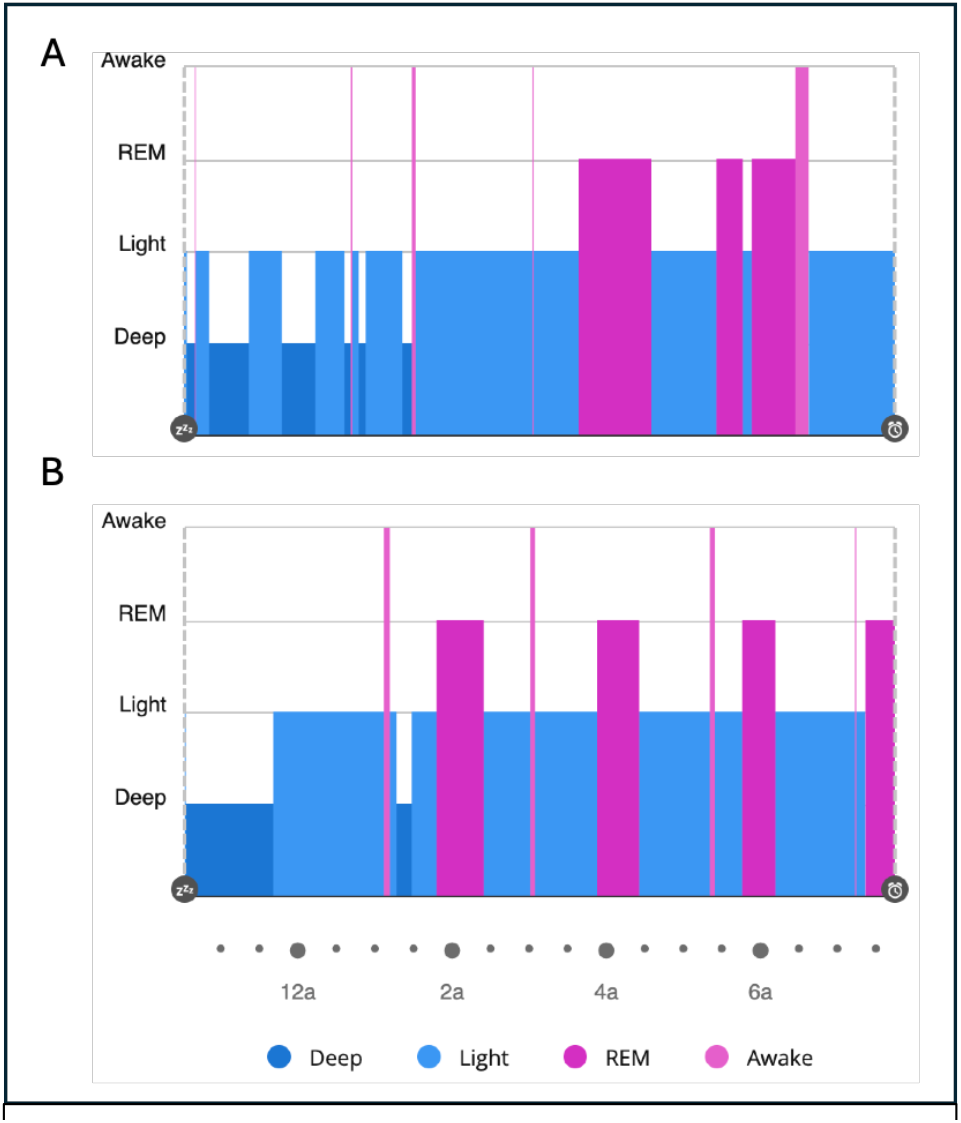
Exemplar Architecture Contrast. Panel A: Feb 25 (AG=2, 78m deep, 6 blocks, longest 31m, OFS 2.9); Panel B: Mar 9 (AG=5, 80m deep, 2 blocks, first 69m, OFS 6.8). Garmin-style sleep timeline visualization for each night with deep=dark, REM=medium, light=light, awake=white. Annotate total deep, architecture group, OFS, environmental conditions.

The N-attempts contextual penalty — applied when five or more distinct deep sleep blocks are detected in a single night — was triggered on n = 16 of 78 clean nights (20.5%; pre-combining block count). On these nights, OPF v5 systematically overestimated OFS (mean residual +0.44 without penalty *versus* +0.12 with penalty), The penalty reduced large misses (residual >0.5 OFS units) by 13% across the clean dataset. Deep:REM ratio on high-N-attempts nights was predominantly below 0.50 (REM-dominant pattern in 14 of 15 penalty-trigger nights), confirming the co-occurrence of architectural fragmentation with homeostatic REM compensation.

### 4.5 Space Weather Correlations

Pearson correlation analysis across three space weather metrics and three temporal alignments (**Table 5**) revealed a dissociation between geomagnetic field disturbance (Kp index) and solar electron flux (daily fluence) as predictors of next-day wellness. Kp Peak achieved significant bivariate correlation with OFS at same-day alignment (r = **−0.249**, p = **0.028**), with no significant effect at 1-day or 2-day lag. Daily electron fluence showed a marginal same-day trend (r = **−0.209**, p = **0.066**, ns) but achieved significance at the 1-day lag (r = **−0.257**, p = **0.024**), consistent with a delayed architectural effect. Load Score was non-significant same-day (r = **−0.192**, p = **0.092**) but showed a significant 1-day lagged association with OFS (r = **−0.328**, p = **0.004**), suggesting that cumulative electron flux burden manifests its functional cost predominantly the following day. Multiple regression of OFS on Architecture Group, Kp Peak, and daily fluence simultaneously achieved R = **0.763**, R² = **0.582** — an improvement of only **1.6%** beyond Architecture Group alone (R² = **0.567**), confirming that space weather adds modest incremental predictive value beyond sleep architecture.

**Table 5.**
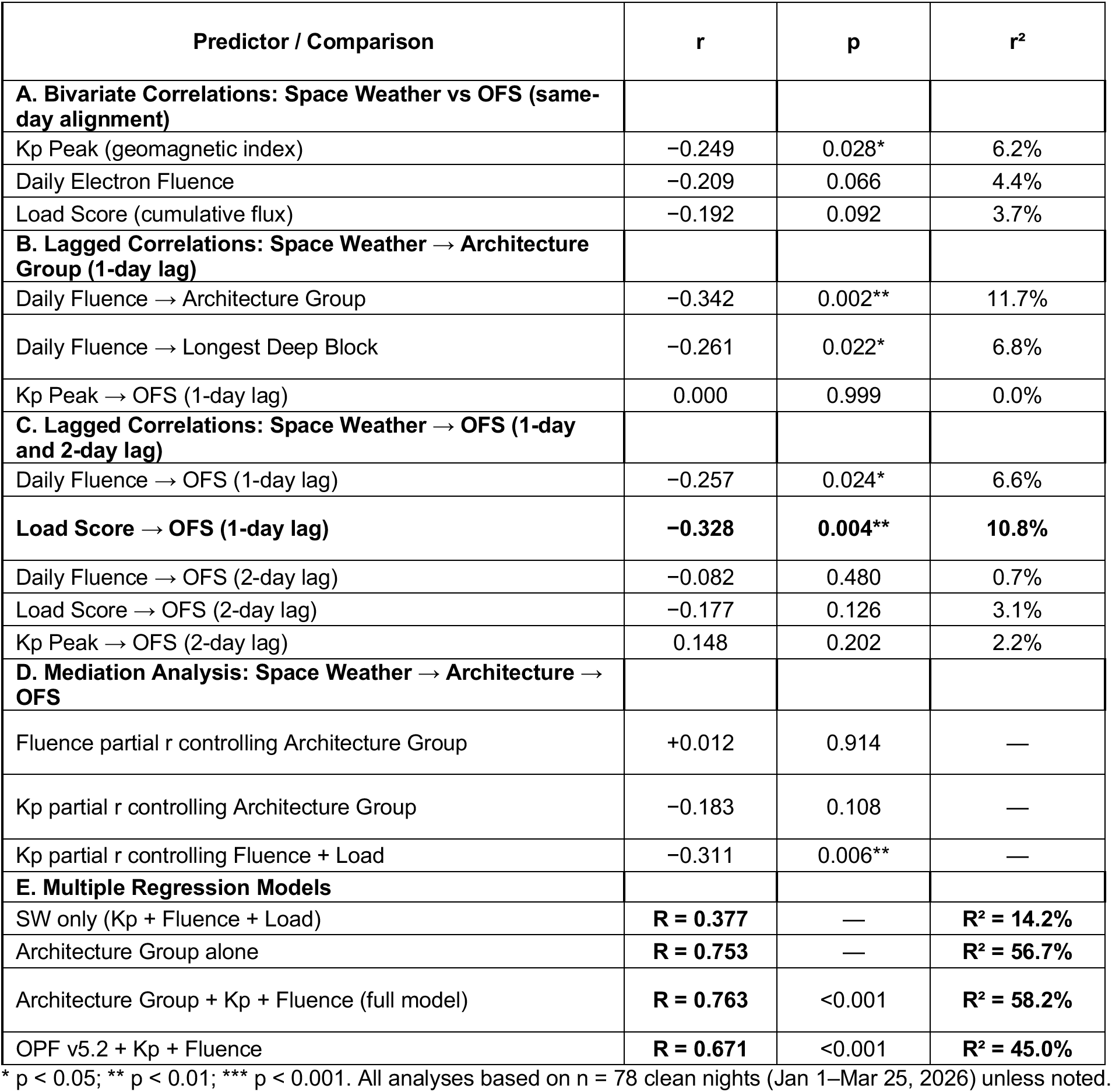
Space Weather Temporal Alignment Analysis and Mediation Results (clean n=78)

Storm timing relative to the deep sleep initiation window further modulated architectural outcomes beyond storm magnitude alone. Geomagnetic storms arriving during the primary SWS window (approximately 10:00 pm–2:00 am) consistently produced greater deep sleep suppression than equivalent-magnitude storms arriving after 3:00 am: a G2 storm (Kp **6.0**) with pre-sleep onset on March 14 produced AG1 with a **32-minute** best deep block despite **77** total deep minutes; a co-occurring G3 storm on March 22 (Kp 7.0) similarly suppressed architecture despite arriving after the primary SWS window; the G3 storm on March 21 (Kp **7.3**, onset 11:28 pm) produced the lowest OFS in the dataset (**0.5**), AG1, and **44** total deep minutes.

### 4.6 Model Performance on Immune/Viral-Confounded Nights

Twenty nights across the full study period — including a viral episode from April 25–May 10 (extended dataset) — were flagged as immune- or viral-confounded and excluded from primary analysis (Table S7; Supplementary Section VII). Post-hoc examination of OPF v5.2 residuals on these nights revealed a systematic pattern: the model overpredicted next-day OFS on 18 of 19 nights (95%), with a mean residual of −0.56 (SD = 0.36, range −1.39 to 0.00). The consistent negative direction of residuals indicates that immune/viral burden reduces OFS through mechanisms the architecture-based model does not capture.

## 5. DISCUSSION

This longitudinal case study provides quantitative evidence that sleep architecture quality, operationalized as the degree to which deep sleep occurs in sustained, consolidated blocks, is a substantially stronger predictor of next-day functional wellbeing than total sleep stage duration in a patient with Long COVID and dysautonomia. Architecture Group (AG) classification explained more than 10 times the variance in OFS compared with Garmin score, and 3 times more than deep sleep time along (**Table 4**). This study documents a central dissociation that has not been characterized in Long COVID using longitudinal wearable biometric data at the individual level [10, 11]: fragmented nights that achieve equivalent or superior total deep sleep duration compared with consolidated nights produce substantially worse next-day function. It extends the established literature on sleep fragmentation in healthy populations [12, 13] into the post-viral dysautonomia context, where the dissociation appears mechanistically amplified: autonomic insufficiency not only increases the probability of fragmented architecture but also reduces the system’s capacity to compensate for fragmentation through other restorative mechanisms.

### 5.1 The Deep Sleep Fragmentation Phenotype: Mechanisms and Consequences

Two mechanistically distinct pathways through which sleep fragmentation impairs restoration were identified and incorporated into the Pendulum model, each validated by their predictive improvement on key miss nights. The first pathway (captured by the 30–34 minute best-block penalty introduced in v5) reflects the non-linear relationship between deep sleep block duration and glymphatic metabolic clearance. Norepinephrine-mediated glymphatic clearance requires sustained N3 epochs to complete biologically meaningful CSF-exchange cycles, with an initiation phase of approximately 10–15 minutes leaving a diminishing effective clearance window in blocks shorter than 40 minutes [14, 15]. The observed performance difference between nights with near-identical total deep sleep yet significantly different longest deep block lengths is consistent with this mechanism (**Figure 3**). The second pathway (captured by the N-attempts penalty applied when five or more distinct deep sleep blocks occur in a single night) reflects the cumulative neuroendocrine cost of repeated failed descent attempts. Each abortive N3 episode is accompanied by a sympathetic micro-activation that, in aggregate, elevates catecholamines, ACTH, and cortisol throughout the 24-hour cycle [16]. The downstream mechanism operates through the CRH axis: as HPA activation accumulates, corticotropin-releasing hormone concentrations reach levels that actively suppress further slow-wave sleep consolidation and diminish the restorative value of compensatory REM [17–20]. This explains the paradox observed on high-N-attempts nights, where quantitatively abundant REM produces less restoration than would be predicted by its duration: the neuroendocrine milieu generated by multiple failed N3 descents impairs REM’s restorative expression.

These two mechanisms are additive but mechanistically distinct: the first operates at the level of the individual block’s biological efficiency; the second operates cumulatively across the night at the neuroendocrine system level. Together they define what we term the ‘*initiation cost hypothesis*’—the observation that deep sleep fragmentation carries physiological costs beyond what stage-duration metrics can capture, and that those costs scale non-linearly with both block brevity and attempt frequency. This concept parallels but extends the cognitive cost framework established in experimental sleep fragmentation studies [12, 13], situating it within a specific glymphatic and HPA-axis mechanistic framework applicable to post-viral dysautonomia. The stutter fragmentation pattern (AG 4), with the highest mean total deep sleep of any group yet OFS outcomes worse than AG5 nights, provides perhaps the most direct evidence that total deep sleep duration is an insufficient metric of restorative value.

### 5.2 The Pendulum Model and Autonomic Gating of Sleep Architecture

The 8.5-fold advantage of OPF v5.2 over the Garmin Sleep Score in explained variance (43.0% vs. 4.8%; **Table 4**) reflects three fundamental limitations of commercial wearable algorithms in this population: population calibration that does not accommodate the atypically low HRV and architectural patterns of dysautonomia; reliance on total stage duration without block-level consolidation weighting; and absence of contextual adjustments for environmental or pharmacological modifiers. The dependence of the Garmin algorithm on overnight HRV as a primary input is particularly problematic here: overnight HRV in this participant averaged 34.3 ms, substantially below expected norms, and is derived from the same sleep period being characterized, introducing circularity that an independent pre-sleep HRV measurement would resolve. v5.2 incorporates biologically motivated refinements identified through systematic residual analysis to address these limitations (**Supplementary Section IV**), with substantial subgroup improvements and no subgroup worsening (**Table S5**). The model’s remaining unexplained variance (57.0% for v5.2) represents genuine unpredictability in autonomic state, unmeasured variables (GI symptoms, psychosocial stressors, nutritional factors), and residual environmental noise. Full cross-validation on an independent dataset remains the appropriate standard for definitive model evaluation.

The data support a model in which ANS state at sleep onset acts as a permissive gate for deep sleep consolidation, necessary but not sufficient, interacting with both homeostatic sleep pressure (Process S) and the circadian regulation of slow-wave sleep expression (Process C; [21, 22]. Within this framework, the timing of autonomic settling relative to the circadian window for slow-wave sleep (roughly the first 90 minutes after sleep onset) is as important as the degree of settling achieved. This explains why overnight HRV, measured as an average across the full sleep period, is a poor predictor of architectural quality in this population: it reflects the integrated autonomic state across all stages rather than the critical pre-consolidation window. ANS state at sleep onset is itself a multidimensional construct that captures sympathovagal balance but not the vestibular, cerebellar, and HPA-axis contributions to sleep stage gating, for which no single proxy metric provides complete characterization [23]. The contextual penalties of the Pendulum model operationalize two of these dimensions directly: the N-attempts penalty captures the cumulative HPA cost of failed gating events across the night, and the saturation modifier captures the carry-forward effect of sustained environmental loading on gate permissivity the following night.

### 5.3 Space Weather as Environmental Modifier: A Two-Pathway Model

One of the more surprising findings in this study was the contribution of space weather to sleep architecture and secondarily to OFS in this patient (**Table 4**). Indeed, the study identified a two-pathway model of space weather effects, in which Kp (geomagnetic field disturbance) acts acutely on autonomic nervous system state and sleep architecture, and electron fluence acts cumulatively through architectural suppression, represents the most novel contribution of this study and the one that requires the most cautious interpretation. The Kp pathway is the better-established of the two. Acute geomagnetic disturbance has been associated with ANS dysregulation, melatonin suppression, circadian disruption, and cardiovascular events in population-level studies [24, 25]. Associations between solar flare, CME, and geomagnetic storm with cardiovascular and autonomic effects have been recently described [26], and suggested to be mediated by melatonin suppression, cryptochrome magnetoreception, and voltage-gated calcium-channel modulation [26, 27]. Mediation analysis in this dataset indicates that Kp operates primarily through architecture degradation rather than through a direct OFS pathway independent of sleep structure (**Table 5**), consistent with the proposed mechanism: geomagnetic disturbance suppresses ANS settling at sleep onset, which in turn prevents consolidated deep sleep, which drives next-day functional decline. Critically, Maghrabi and Maghrabi [26] characterize neurological and psychological space weather associations as ‘preliminary’, a framing the findings of this study support and extend into the sleep architecture domain. The storm timing finding documented here shows that geomagnetic storms that begin during the primary slow wave sleep window from 10 pm to 2 am produce greater disruption of sleep architecture than storms of similar intensity that begin after 3 am. This pattern is mechanistically consistent with the autonomic nervous system gating model. A storm that occurs after the primary period of consolidated deep sleep cannot undo the restoration that has already taken place, whereas a storm at sleep onset can prevent the first consolidated sleep cycle from occurring. To our knowledge, this temporal dependence has not been previously reported.

The electron fluence pathway is more complex and remains tentative. Fluence showed a marginal same day association with sleep outcomes, but it predicted Architecture Group at a one-day lag (**Table 5A**). This relationship was fully mediated by sleep architecture, as the partial correlation controlling for Architecture Group was essentially zero (**Table 5D**). This delayed effect, which operates entirely through degradation of sleep architecture, differs from the acute mechanisms proposed for Kp, such as melatonin suppression and magnetoreception. Instead, it points to a distinct cumulative process. One possibility is that sustained elevation of relativistic electron flux maintains a background state of cortical hyperexcitability. This could occur through magnetoreception pathways, which may interfere with the generation of synchronized delta waves without producing the acute cardiovascular and autonomic symptoms typically associated with geomagnetic storms [26]. The observed negative correlation between Kp and electron fluence (**Table 5C**) further supports treating them as distinct heliophysical phenomena that tend to occur during different phases of solar activity. This distinction is important because it indicates that future studies of space weather and sleep should not treat different space weather metrics as interchangeable. A striking finding from the lagged correlation analysis warrants particular attention. Load Score at a one-day lag emerged as the strongest single space weather predictor in the dataset (**Table 5C**), exceeding the effect size of same day Kp Peak (**Table 5A**). This temporal separation is consistent with a biological mechanism operating on a longer timescale than acute autonomic nervous system perturbation.

### 5.4. Architecture as a Leading Indicator of Functional Recovery

In the extended monitoring period (**Table S3**), a pattern consistent with hysteresis was observed following a sustained elevated Load Score run: Architecture Group remained predominantly AG1 throughout, recovering to AG3–5 only after Load Score stabilized for three or more consecutive nights. This delayed recovery is consistent with three features of the proposed space weather model: (1) a system that recovers more slowly than it activates; (2) HPA axis sensitization under repeated sub-threshold electromagnetic stress; and (3) a fundamental difference in temporal dynamics between stressors that accumulate gradually (electron fluence, Load Score) and those that arrive as discrete impulses (geomagnetic storms, Kp index), with the former producing delayed and persistent effects.

The same asymmetry, characterized by rapid degradation and slow recovery, was observed across heterogeneous triggers including intercurrent viral illness and pharmacological disruption, not only space weather loading (**Table S3; Table 3**; see also Section 5.5 for evidence that viral illness additionally reduces OFS through architecture-independent pathways). On 8 of 9 first-recovery nights across the full monitoring period, OFS fell meaningfully below the mean typically associated with that architecture group, suggesting that architecture quality functions as a leading indicator of functional recovery rather than a concurrent one. A single consolidated night after a degraded run signals that recovery has begun, not that it has arrived. These findings should be interpreted cautiously given the single-patient design and likely heightened environmental sensitivity associated with dysautonomia. Prospective studies in larger cohorts are needed to determine whether the effect sizes observed here generalize. This perspective aligns with Maghrabi and Maghrabi [26], who described geomagnetic disturbances as a novel environmental risk factor for vulnerable patients; the present study adds two specific contributions, identifying sleep architecture as the plausible mechanism and proposing a temporal gating model that specifies when geomagnetic events are most likely to cause disruption.

### 5.5 Methodological Considerations and Limitations

This study employs a prospective N-of-1 longitudinal design, enabling within-patient physiological inference at a resolution not achievable by group-level studies, while accepting that external validity is limited to the individual studied [9] Wearable-derived sleep staging introduces inherent limitations for N3 detection: PPG and accelerometry-based algorithms do not resolve sleep stages at PSG resolution, and the device has not been validated in dysautonomia populations. Absolute deep sleep minute values should be interpreted as approximations. The Architecture Group classification, based on block-level structure rather than aggregate N3 minutes, is less sensitive to absolute staging accuracy and more robust to proportional errors that preserve block structure; within-patient correlational analyses are similarly robust to systematic staging error that is stable across nights. The Overall Feeling Score is a subjective outcome reported each morning without prior knowledge of the night’s biometric data, establishing assessor independence, but it is not a validated clinical instrument; standardization against validated instruments (e.g., PROMIS Fatigue, EQ-5D) would strengthen interpretation. The patient-researcher identity creates potential demand characteristics that cannot be fully excluded, though the morning-first, biometric-blind reporting protocol mitigates this concern.

Several additional limitations are acknowledged. First, all findings derive from a single patient; replication in additional PASC patients with dysautonomia is required before generalization. Second, the absence of concurrent PSG prevents confirmation of architecture classifications and arousal index data. Third, space weather correlational analyses involve multiple comparisons; of the metrics examined, only Load Score 1-day lag survives conservative Bonferroni adjustment (**Table 5**). Fourth, mediation analysis cannot establish causality; the Kp and fluence pathway models are structural models consistent with the data, but causal inference requires experimental manipulation not available in an observational design. Fifth, the iterative model development approach introduces circularity; the post-dental-device monitoring period beginning April 27, 2026 constitutes the first out-of-sample prospective test of the model and will be reported separately.

Sixth, immune and viral flares represent an unmeasured confound not addressed by the current model (**Supplementary Section VII; Table S7**). These findings are consistent with the hysteresis framework in Section 5.4: illness degrades architecture through the same pathway as space weather loading, but the residual signal reflects a direct neuroinflammatory burden on autonomic tone, cognitive function, and symptom capacity that the architecture-based model cannot capture, particularly consequential in Long COVID dysautonomia, where baseline ANS reserve is already compromised.

### 5.6 Conclusions

This study demonstrates that in a patient with Long COVID and dysautonomia, deep sleep architecture quality characterized by block consolidation, first-cycle depth, and absence of fragmentation, is a substantially stronger predictor of next-day functional wellbeing than total deep sleep duration (**Table 4**). The deep sleep fragmentation phenotype, present on 25.6% of clean nights, is associated with poor outcomes despite adequate total deep sleep and is mechanistically attributed to incomplete glymphatic clearance cycles and cumulative HPA-axis activation from repeated failed descent attempts (i.e., the initiation cost hypothesis).

An important question for future investigation is how vulnerable dysautonomia patients are to environmental stressors, whether electromagnetic, infectious, or otherwise. Once a patient establishes a stable clean protocol (**Supplementary Section Ia**), real-world disruptions can crash sleep architecture rapidly and prove resistant to equally rapid recovery. This hysteresis dynamic was observed across heterogeneous trigger types in this dataset (**Section 5.4**) and is consistent with a system in which functional recovery lags structural sleep recovery. Similarly, the proportion of illness-flagged nights and the extended duration of the April–May viral episode are consistent with the prolonged immune dysregulation described in Long COVID.

Patients with Long COVID dysautonomia may be particularly vulnerable to immune and viral flares [28], which appear to reduce next-day functional capacity through pathways that operate partly independently of sleep architecture disruption. Direct neuroinflammatory effects on autonomic tone, cognitive function, and symptom burden are not captured by an architecture-based model such as OPF v5.2, and immune-confounded nights were accordingly excluded from primary analysis. Nevertheless, the n=1 longitudinal design affords a method for quantifying this contribution: by applying the architecture-based model to flagged immune nights and examining residuals, it is possible to isolate the non-architecture component of immune burden (**Section 4.6**). This suggests that n=1 longitudinal models in Long COVID and the methodological framework described here may serve not only as predictive tools, but as instruments for parsing the independent contributions of sleep, immune state, and environmental load to day-to-day functional capacity.

The methodological framework described here may serve as a hypothesis generator for group-level studies in Long COVID sleep research. The block-level fragmentation phenotype bears structural resemblance to architecture disruption described in other conditions where ANS or neurological dysregulation impairs sleep stage consolidation, including SCN8A-related developmental and epileptic encephalopathy [29], and may be applicable across post-viral and autonomic disorders more broadly.

## DECLARATIONS

### Ethics Approval and Consent to Participate

This study was conducted in accordance with the Declaration of Helsinki. The study protocol involving a N-of-1 design and a single participant was reviewed by the Institutional Review Board of the University of Arizona and determined that the proposed activity is not research involving human subjects as defined by DHHS and FDA regulations [IRB protocol number: STUDY00008366; approval/exemption date: June 25,2026].

### Consent for Publication

Written informed consent for publication of this case report and any accompanying data was obtained from the study participant. As the participant is also the corresponding author, consent for publication is inherent. A copy of the consent documentation is available to the Editor-in-Chief upon request.

### Availability of Data and Materials

Data sufficient to reproduce the primary analyses reported in this study are available in the Supplementary Material, which includes nightly summary statistics, architecture classification distributions, model performance comparisons, space weather correlations, and the illness residual analysis (Tables S1–S7). Additional de-identified data underlying the analyses (sleep architecture variables, space weather metrics, Overall Feeling Score, and OPF model outputs) are available from the corresponding author upon reasonable request. Space weather data (Kp index, GOES 2 MeV electron flux) are publicly available through the National Oceanic and Atmospheric Administration Space Weather Prediction Center (https://www.swpc.noaa.gov).

Raw Garmin device data are subject to the Garmin Connect Terms of Service and cannot be redistributed.

### Authors’ Contributions

Contributions are described using the CRediT (Contributor Roles Taxonomy) framework. MFH: Conceptualization, Data Curation, Formal Analysis, Investigation, Methodology, Project Administration, Software (OPF model development), Visualization, Writing – Original Draft, Writing – Review & Editing. SP: Clinical Supervision, Validation (clinical interpretation), Writing – Review & Editing, IRB oversight. The author read and approved the final manuscript.

## Acknowledgements

I am extremely grateful to Sairam Parthasarathy, MD for inspiration and guidance. The authors thank the National Oceanic and Atmospheric Administration Space Weather Prediction Center for maintaining publicly accessible geomagnetic and electron flux data used in this study. The authors also acknowledge the BIO5 Institute at the University of Arizona for institutional support.

## Authors’ Information (optional)

Michael F. Hammer, PhD, is a Research Scientist at the BIO5 Institute, University of Arizona, Tucson, AZ. His research interests include longitudinal physiological monitoring and quantitative modeling of biological systems.

